# Radiation Free Percutaneous Nephrolithotomy (PCNL) is *Not* Always Feasible *comparative prospective trial*

**DOI:** 10.1101/2025.07.30.25332444

**Authors:** Noah Swärd, Anthony Galvez, Riley Hull, Joseph Girgiss, Jonathan Katz, Jake L. Roberts, Tyler J. Sheetz, Luke K. Griffiths, Martin Ugander, Roger L. Sur

## Abstract

**Introduction and Objective:** Percutaneous nephrolithotomy (PCNL) is a minimally invasive procedure for kidney stone removal traditionally guided by fluoroscopy. This study aimed to evaluate the feasibility and outcomes of radiation-free PCNL using ultrasound alone compared to standard fluoroscopy-guided PCNL.

**Methods:** A total of 63 PCNL cases were eligible for radiation-free PCNL, but 27 were excluded (intra-operatively aborted, ureteroscopy performed instead, pre-operative complex anatomy). Of the remaining 36 cases eligible for radiation-free PCNL, 11 were converted intra-operatively to fluoroscopic based PCNL. Post-operative CT imaging was available for only 16 of the 25 radiation-free PCNL cases and 4 of the 11 converted cases. We designated these 16 prospective radiation-free PCNL cases (2024–2025) as Group A. For comparison purposes we identified a historical case-matched cohort of 150 PCNL’s. Of these 67 were excluded (similar reasons) leaving 83 retrospective fluoroscopy-guided PCNL cases (2022–2024) called Group B. The *primary outcome* was stone-free rate (SFR), assessed post-operatively by non-contrast CT (2–3 mm slices). *Secondary outcomes* included estimated blood loss (EBL), complication rates (Clavien-Dindo), and post-operative stone events.

**Results:** The median pre-operative stone burden was 35 mm in Group A and 27 mm in Group B and C [p=0.3]. SFR (Grade A) was comparable across Groups A, B, and C [38%, 30%, 25% respectively (p = 0.8)]. No differences were observed in complications or secondary outcomes.

**Conclusions:** Radiation-free PCNL is feasible and yields comparable outcomes to standard fluoroscopy-guided PCNL, offering a promising method to reduce radiation exposure without compromising surgical success. However, we identified a consistent theme of poor visualization that led to large proportion of cases requiring conversion to fluoroscopy. Innovation directed towards improving tool echogenicity is key to diffusing this technique.

## Introduction

Fluoroscopy-guided percutaneous nephrolithotomy (PCNL) remains the standard of care for removing renal stones larger than 20 mm ^1^. Traditionally, fluoroscopy is used during multiple steps of the procedure, including needle access, guidewire manipulation, tract dilation, and assessment of residual stones at the conclusion of the case ^2^. However, fluoroscopy exposes patients and providers to ionizing radiation, which carries both deterministic and stochastic risks. Transitioning to radiation-free imaging modalities during PCNL is therefore of significant clinical value.

Ultrasonography has been proposed as a viable alternative to fluoroscopy for various steps of PCNL, with particular benefit in reducing radiation exposure among vulnerable populations such as pregnant women, pediatric patients, and individuals with renal impairment ^3^. Although concerns exist regarding reduced imaging quality and surgical accuracy, some studies have shown that ultrasound can improve precision during needle access by providing real-time, three-dimensional anatomical visualization ^4^. Recently, fully ultrasound-guided PCNL has been explored, yielding promising results ^5^.

While similar comparative studies exist, ultrasound-guided PCNL remains highly operator-dependent, with outcomes influenced by surgeon experience and technique. Moreover, all prior literature compares “successfully” performed ultrasound only PCNL to fluoroscopic PCNL. There is little discussion of why ultrasound PCNL attempt fails at times. As such, it is important for individual institutions to evaluate their capacity to adopt a radiation-free approach. This study aims to assess the feasibility of using ultrasound as the <u>sole imaging modality throughout the entire PCNL procedure</u>. The results of this prospective cohort study may contribute to evolving standards for radiation-free stone surgery.

## Methods

This prospective cohort study compared subjects undergoing complete radiation-free PCNL (Group A) versus a retrospective cohort of subjects who underwent fluoroscopic guided (Group B). The study was approved by the institutional review board (protocol #180946) and registered at ClinicalTrials.gov (NCT06922006). From March 2024 a single fellowship-trained urologist (RLS) began a systematic attempt to perform all PCNL cases using a radiation-free technique ^6^. The urologist had 20 years of PCNL experience and had routinely used ultrasound assisted renal access since 2015, but never attempted a radiation free PCNL. Group A included patients who underwent radiation-free PCNL between March 2024 and June 2025, where fluoroscopy was not intended. Group B was a case-matched retrospective cohort of fluoroscopy-guided PCNLs performed by the same surgeon between 2021 and 2024, prior to the implementation of radiation-free PCNL. Cases from Group A that were converted intra-operatively to fluoroscopy were labeled Group C.

To maintain patient safety, fluoroscopy equipment was available in all cases and used only if ultrasound imaging was deemed insufficient. Of note, it is the practice of this surgeon to consent patients for endoscopic combined retrograde PCNL--with the understanding that PCNL might not be necessary if retrograde approach sufficient to treat stone burden. All data were stored in REDCap®, a secure, HIPAA-compliant online database. Categorized into demographics, pre-operative data, operative data and post-operative data.

The *primary outcome* was proportion of stone-free, stone-free rates (SFR), classified into three grades based on non-contrast CT scans with 2–3 mm cuts performed postoperatively: Grade A: No residual stones; Grade B: Grade A + ≤2 mm fragments; Grade C: Grade B + 2.1–4 mm fragments*. Secondary outcomes* of this study included hospital length of stay, post-operative stone size, 30 days post-operative events (readmissions, unplanned events, unplanned procedures, emergency department visits), and complications.

### Surgical technique and preparation

Except for the imaging modality, all PCNLs followed standard surgical protocols, in accordance with what has previously been suggested in existing literature about radiation free PCNL^6^. Patients were positioned supine or prone based on surgeon discretion, with supine preferred unless preoperative clinic ultrasonography suggested poor visualization (unobstructed view of kidney). Supine positioning placed both legs in lithotomy. Both ipsilateral flank and external genitalia were prepped. For prone positioning, both ipsilateral back and external genitalia were prepped. Flexible cystoscope and retrograde wire were placed followed by ureteral access sheath (11/13Fr or 12/14Fr). No fluoroscopy was used during retrograde procedures. A flexible ureteroscope was then passed retrograde to assess stone burden and potential access. Stone was treated in situ with holmium laser lithotripsy if anatomy/stone burden deemed favorable. This retrograde option would occasionally obviate the need for PCNL treatment.

BK 5000 Ultrasound with 1-5mHz 5C1 curved array transducer ultrasound probe (Burlington, MA) with needle guide was placed over flank or back, for supine or prone, respectively. An 18 gauge Boston Scientific Naviguide needle (Marlborough, MA) was then used to identify an optimal calyx. The skin to calyx distance was measured. All dilators were then pre-measured and marked with this known distance to facilitate correct depth placement (Figure 1): 6Fr 20cm Cook Teflon dilator (Indianapolis, IN), 8Fr, 10Fr, 12Fr Boston Scientific Amplatz dilators, 28cm 12/14 Boston Scientific Naviguide access sheath (Marlborough, MA).

**Figure 1.**
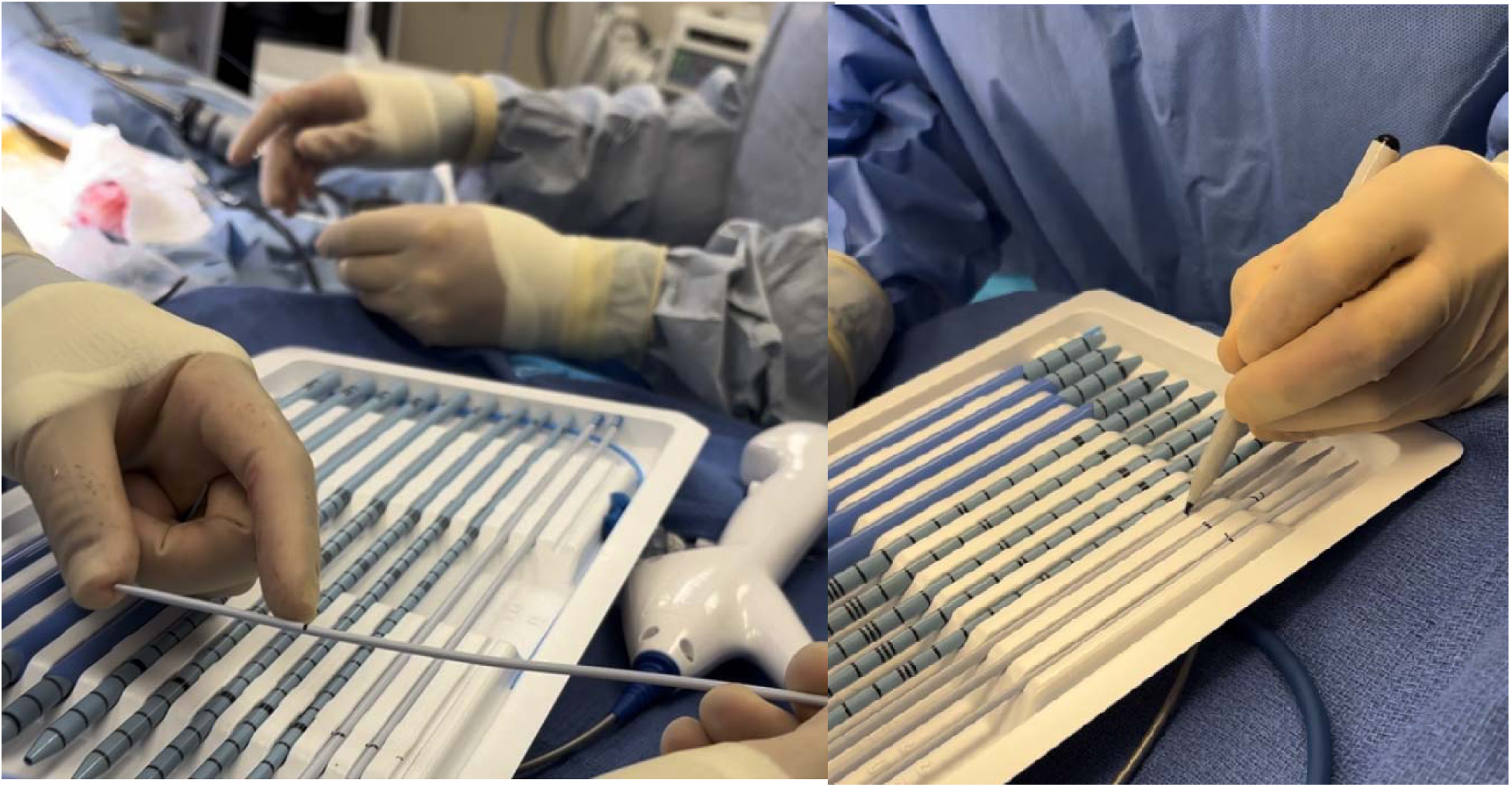
Measuring the dilators.

After successful needle placement a Boston Scientific Amplatz Super Stiff guidewire (Marlborough, MA) was placed into the collecting system. This wire was chosen because of its highly echogenic visibility vice other commercially available wires (Figure 2). Successive dilation of the tract with the above pre-marked dilators was performed under ultrasonography. The correct depth placement was confirmed by both visualizing the dilator marking at the skin as well as visualizing the disappearance of Amplatz Super Stiff guidewire under ultrasonography (Figure 3). The guidewire disappeared under ultrasound since the dilators were not echogenic and their passage over the guidewire “hid” the guidewire. After sequential placement of the 6Fr, 8Fr, 10Fr and 12Fr, the 12/14Fr access sheath was passed into the collecting again using both the marking and “disappearance” of the guidewire to ensure correct depth placement. Through the access sheath a flexible ureteroscope confirmed collecting system visualization. And then a second guidewire was passed down the ureter through the working channel of the ureteroscope. The access sheath was removed and replaced with a pre-marked 24Fr Boston Scientific Nephromax balloon (Marlborough, MA) using the above techniques to ensure correct depth. The balloon was dilated and it could be seen under ultrasonography. Over this a pre-marked sheath was passed to the same depth. The balloon was removed and standard lithotripsy is performed with the Boston Scientific Trilogy (Marlborough, MA). The end of the case occurred after visual confirmation of stone free status. Retrograde stent was placed, and its correct position was confirmed with flexible cystoscopy. Anticoagulation management followed institutional guidelines: DOACs held for 48 hours before surgery, clopidogrel was discontinued 7 days prior, but aspirin 81 mg was not discontinued.

**Figure 2.**
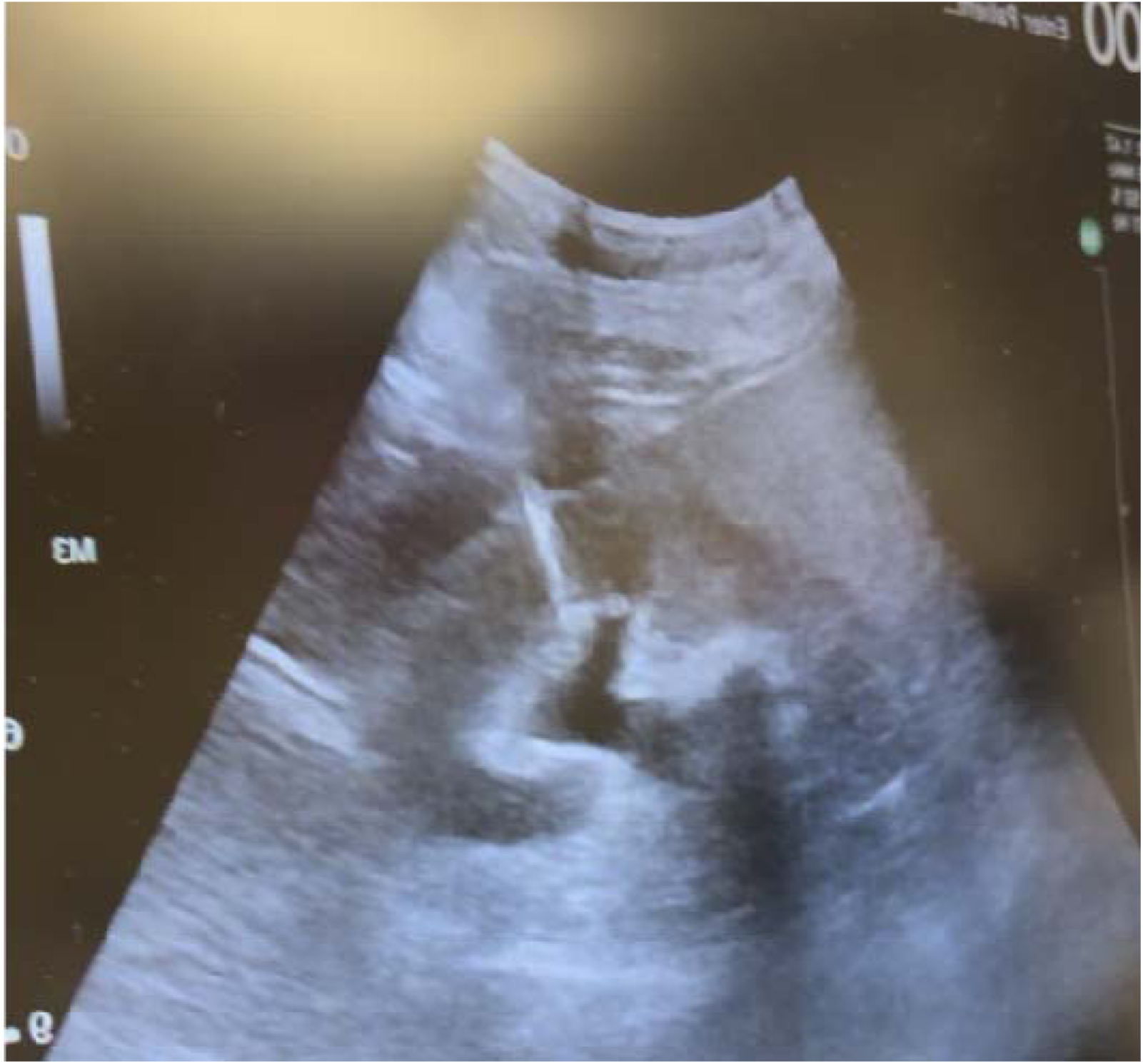
The super stiff wire (very echogenic).

**Figure 3.**
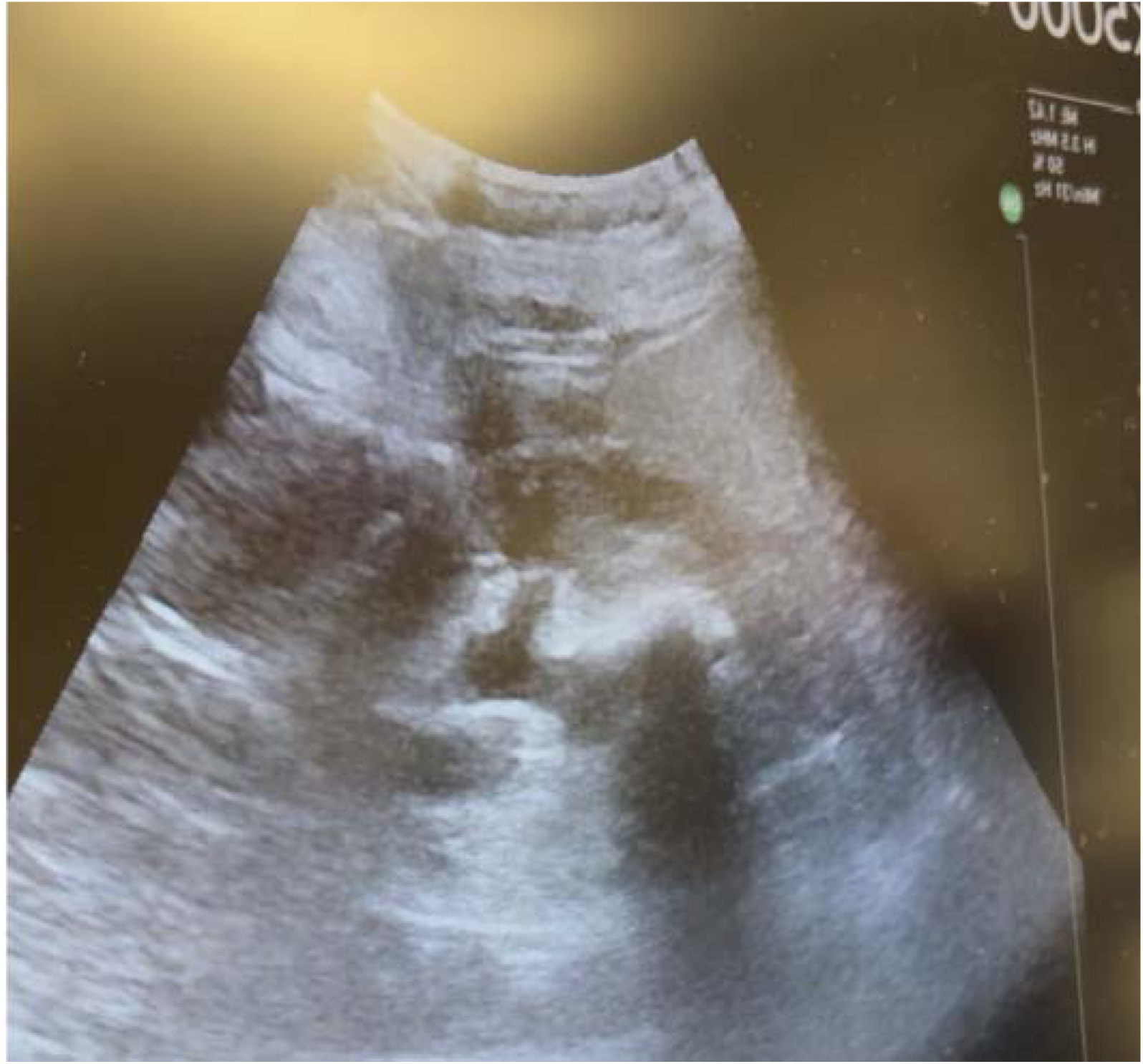
Disappearance of the wire as a dilator is passed over it.

### Statistics

Statistical analyses were performed using Jamovi. Chi-square tests were used for categorical variables and ANOVA for continuous variables. ^7,8^. Bivariate analyses assessed potential confounders of the primary outcome. A p-value < 0.05 was considered statistically significant.

Normality and homogeneity were assessed using the Shapiro-Wilk and Levene’s tests. Since all variables violated assumptions of normality or homogeneity, non-parametric tests were used. Post hoc analysis was performed on one variable that had p<0.05, using the Dwass-Steel-Critchlow-Fligner pairwise comparison.

## Results

### Prospective Cohort

Sixty-three subjects were eligible for radiation-free PCNL (Group A), but 7 cases had to be aborted (Table 5). Seven cases were intraoperatively excluded since retrograde ureteroscopy sufficiently treated the stone burden prior to any percutaneous procedure. Thirteen cases were unsuitable to undergo radiation-free PCNL due to complicated anatomy (n=5), second look PCNL (n=4) and for other reasons (n=4). Thirty-six subjects therefore were planned for radiation-free PCNL. However, only 25 subjects were completed with the radiation-free protocol as 11 subjects (Group C) were converted to radiation PCNL. The reasons for conversion were to verify anatomy (n=5), improve visualization (n=3), verify operative urinary drainage (n=2) and inability to access (n=1). Nine cases from the radiation free group and 7 from the converted group did not have post operative non contrast CT scan with 2-3mm cuts and were excluded—which respectively left 16 and 4 analyzable for the stone free rate, respectively (Figure 4).

**Figure 4.**
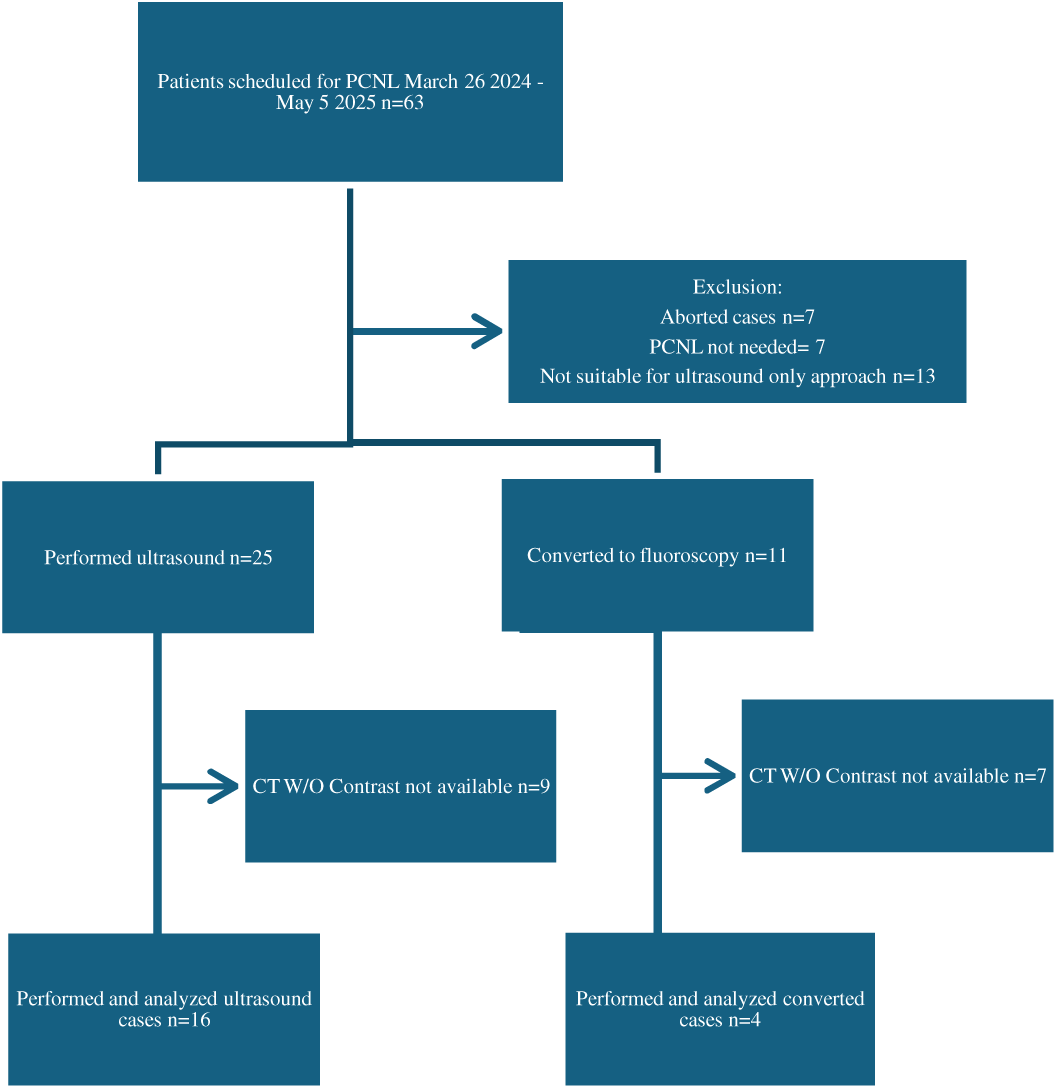
Patient flow diagram showing sample size and group division for the prospective radiation free cohort undergoing PCNL with ultrasound.

### Retrospective Cohort

One-hundred-fifty case-matched PCNL (Cohort B) were analyzed but 6 subjects were aborted (Figure 5). Four cases did not have available pre-operative data. For 15 cases it was determined that PCNL was not needed since their stones were removed ureteroscopically—i.e. we did not need to perform PCNL at this point. Forty-two cases did not have post-operative non contrast CT scan with 2-3mm cuts. This left 83 subjects planned for the comparative fluoroscopic PCNL cohort.

**Figure 5.**
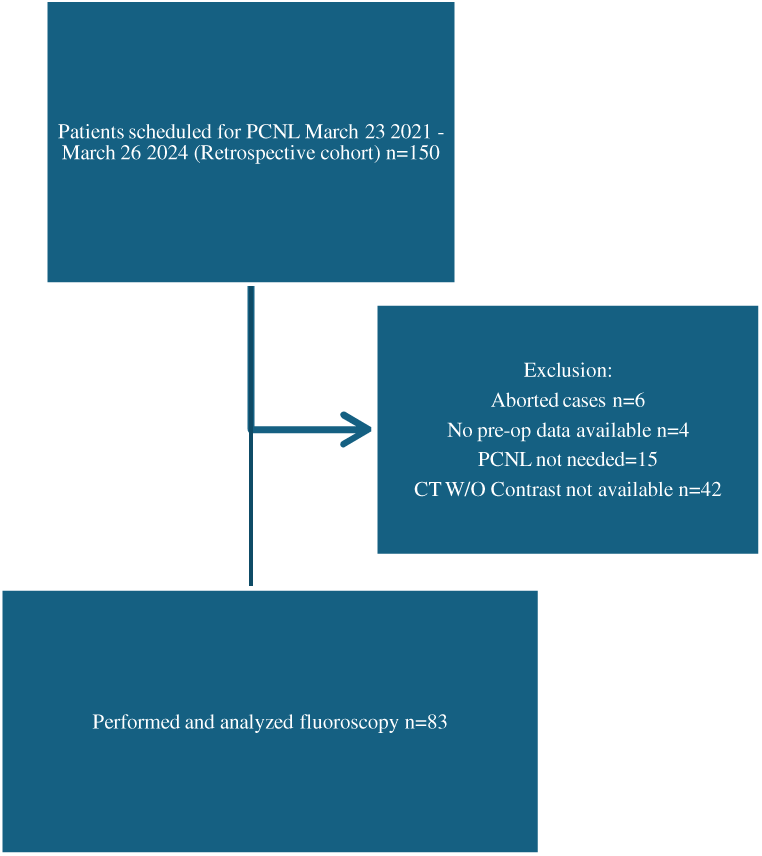
Patient flow diagram showing sample size and group division for the retrospective cohort undergoing fluorosco guided PCNL.

### Patient demographics, pre-operative and operative variables

Patient demographics were similar between the three groups except for an observed difference in ethnicity [p<0.001]. Operative characteristics were similar except for observed differences in patient positioning [p<0.001], tract dilation size [<0.01] and the use of Trilogy lithotripter for removal of stone [p<0.05].

### Primary and secondary outcomes

Regardless of definition (Grade A, B or C), SFR did not differ between procedures done with ultrasound (Group A) compared to fluoroscopy (Group B) (Figure 6). Also, there was no difference in SFR for radiation-free cohort (Group A) compared to SFR for those converted to radiation (Group C). Similarly, there was no difference in SFR for radiation cohort (Group B) compared to those converted to radiation (Group C). Lastly, no differences were seen for all secondary endpoints among the three cohorts (Table 3).

**Figure 6.**
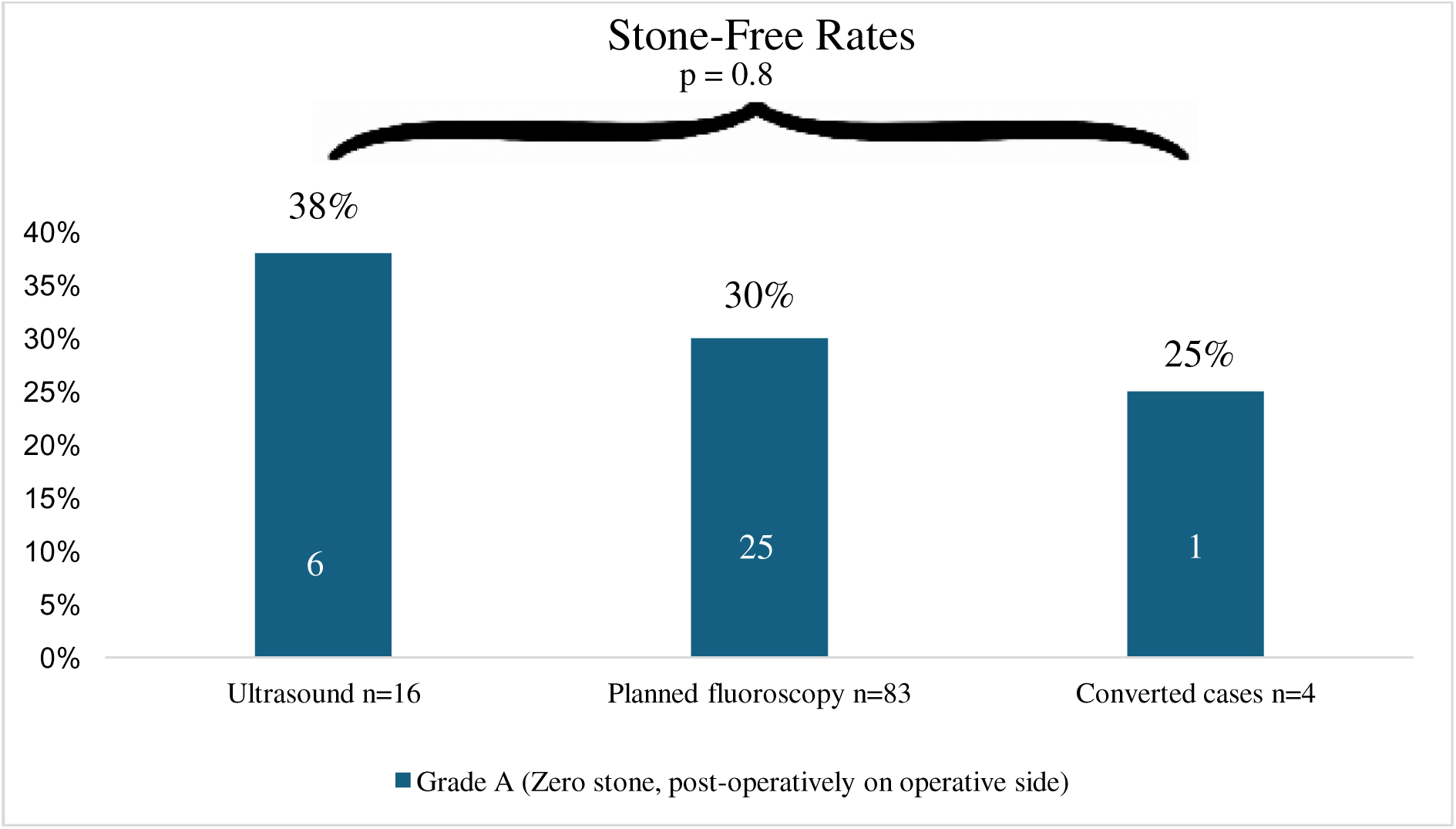
SFR (Grade A, zero stone) were not significantly different among all three cohorts, p=0.8.

**Table 1.**
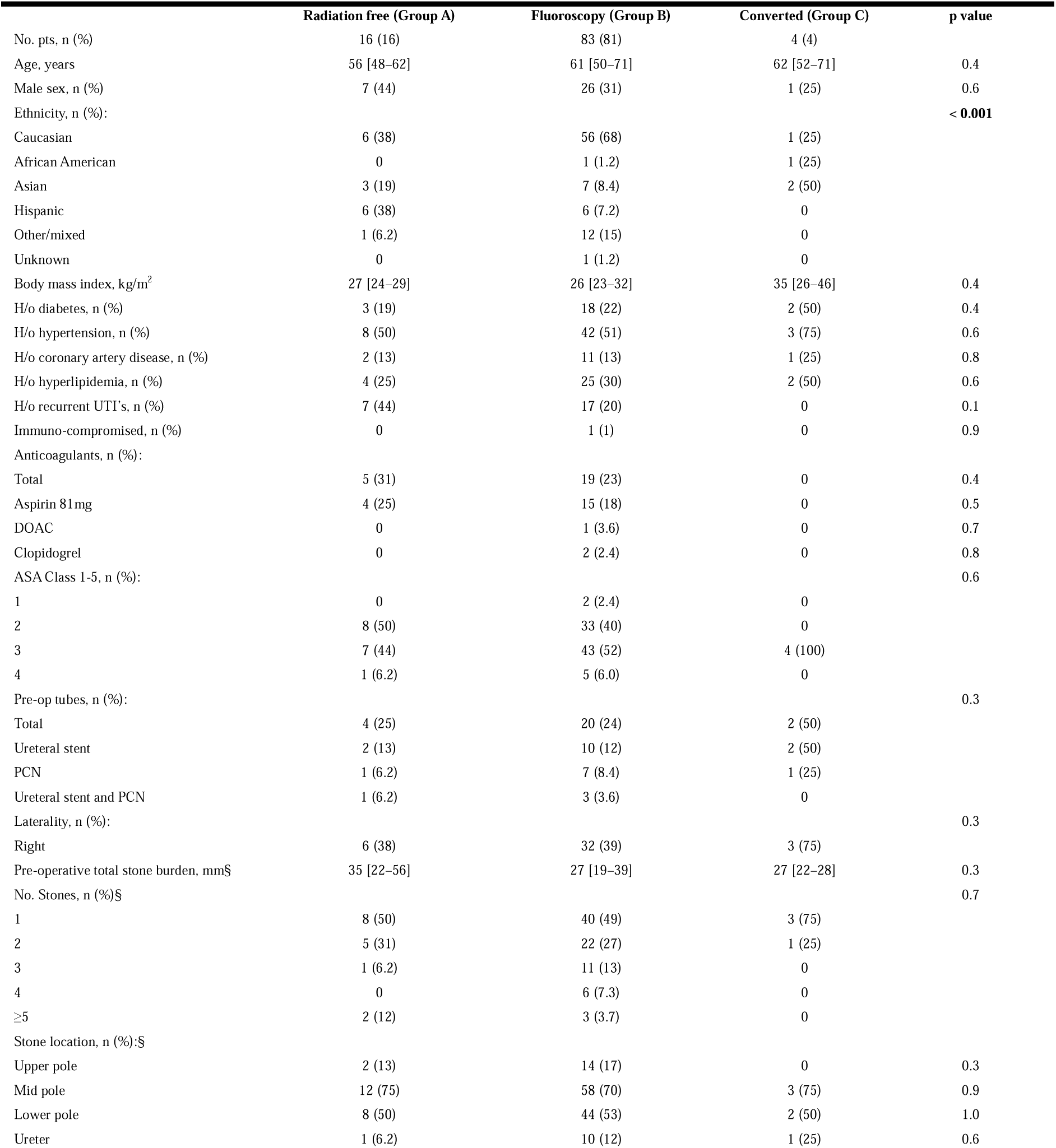
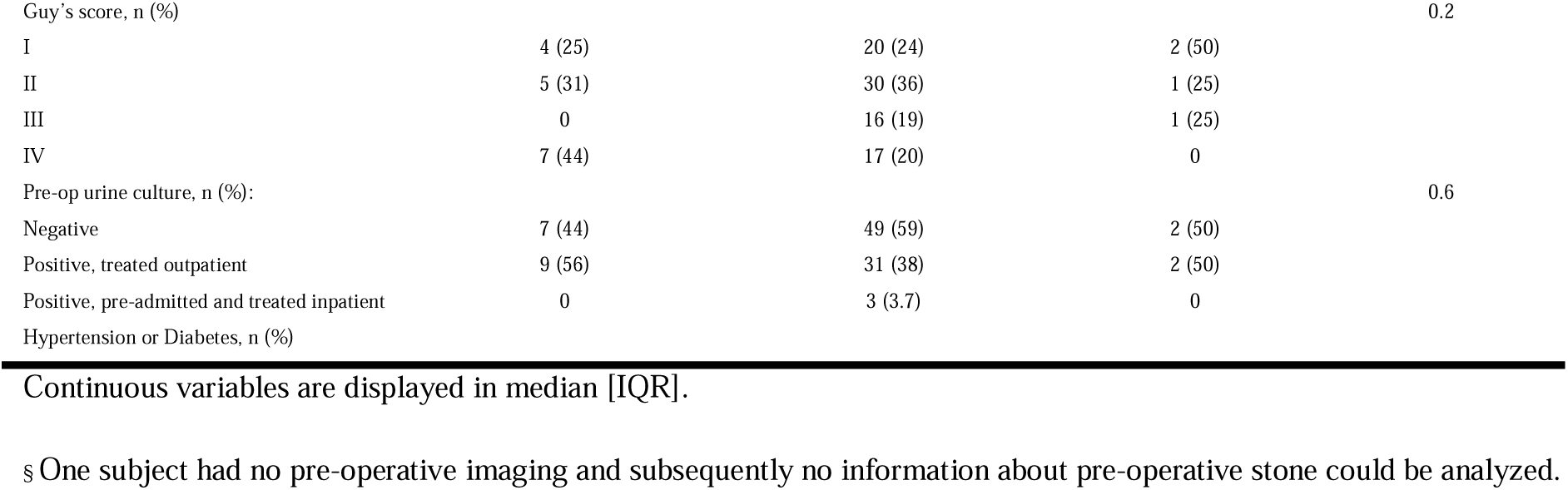
Patient demographics and pre-operative data.

**Table 2.** Operative and post operative data.

|  | Radiation free (Group A) | Fluoroscopy (Group B) | Converted (Group C) | p value |
| --- | --- | --- | --- | --- |
| No. pts, n (%) | 16 (16) | 83 (81) | 4 (4) |  |
| Procedure done in prone position, n (%) § | 2 (12) | 62 (75) | 3 (75) | **<0.001** |
| Access location, n (%): |  |  |  |  |
| Lower pole | 9 (56) | 47 (57) | 2 (50) | 1.0 |
| Mid pole | 6 (38) | 11 (13) | 1 (25) | 0.1 |
| Upper pole | 1 (6.2) | 22 (27) | 1 (25) | 0.2 |
| Indwelling PCN | 0 | 3 (3.6) | 0 | 0.7 |
| No. radiation free access attempts, no (%): §§ |  |  |  | 0.6 |
| 1 | 13 (81) | 41 (77) | 3 (75) |  |
| 2 | 1 (6.2) | 7 (13) | 0 (0) |  |
| 3 | 2 (12) | 2 (3.8) | 1 (25) |  |
| 4 | 0 (0) | 1 (1.9) | 0 (0) |  |
| Tract dilation size, n (%) |  |  |  | **<0.01** |
| 30 Fr | 4 (25) | 30 (36) | 0 (0) |  |
| 24 Fr | 6 (38) | 15 (18) | 4 (100) |  |
| 17.5 Fr | 6 (38) | 38 (46) | 0 (0) |  |
| Stone treatment modality, n (%): |  |  |  |  |
| Trilogy | 12 (75) | 42 (51) | 4 (100) | **<0.05** |
| Holmium laser lithotripsy | 8 (50) | 40 (48) | 0 (0) | 0.2 |
| Perc-n-circle | 2 (12) | 3 (3.6) | 0 (0) | 0.2 |
| Thulium Fiber Laser | 0 | 1 (1.2) | 0 | 0.9 |
| End of case tube, n (%): |  |  |  | 0.1 |
| PCN | 1 (6.2) | 23 (28) | 0 |  |
| Stent | 15 (94) | 60 (72) | 4 (100) |  |
| Total time under fluoroscopy, seconds §§§ | N/A | 107 [75–172] | 105 [79–147] | 1.0 |
| Total cumulative radiation dose, mGy §§§ | N/A | 19 [11–39] | 50 [35–80] | 0.2 |
| Same day discharge, n (%) | 2 (13) | 5 (15) | 2 (15) | 1.0 |
| Time until follow up CT, days | 44 [20–55] | 50 [19–79] | 35 [3–98] | 0.7 |

Continuous variables are displayed in median [IQR]
§ All procedures were performed in prone or supine positioning.
§§ Not all cases had radiation free attempts.
§§§No information about time under fluoroscopy in 1 subject from converted group. No information about total time under fluoroscopy and total radiation dose mGy in 1 additional subject.

**Table 3.** Primary and secondary outcomes.

|  | Radiation free (Group A) | Fluoroscopy (Group B) | Converted (Group C) | p value |
| --- | --- | --- | --- | --- |
| No. pts | 16 (16) | 83 (81) | 4 (4) |  |
| Same day discharge, n (%): | 2 (13) | 5 (15) | 2 (15) | 1.0 |
| Discharge antibiotics, n (%): | 8 (53) | 10 (30) | 5 (38) | 0.3 |
| Time until follow up CT, days | 44 [20–55] | 50 [19–79] | 35 [3–98] | 0.7 |
| Grade A (Zero stone) n (%): | 6 (38) | 25 (30) | 1 (25) | 0.8 |
| Grade B (Stone $\leq 2$mm) n (%): | 7 (44) | 26 (31) | 1 (25) | 0.6 |
| Grade C ($\leq 4$mm) n (%): | 10 (63) | 35 (42) | 1 (25) | 0.2 |
| >4mm stone (Not stone free), n (%): | 6 (37) | 48 (58) | 3 (75) | 0.2 |
| Hospital length of stay, days | 2 [2–2] | 2 [2–3] | 2 [2–2] | 0.4 |
| Issues within 30 days, n (%): |  |  |  |  |
| No issue | 14 (88) | 63 (76) | 2 (50) | 0.3 |
| Re-admission | 1 (6.2) | 12 (14) | 2 (50) | 0.1 |
| ED visit | 2 (12) | 11 (13) | 1 (25) | 0.8 |
| Requires additional stone treatment, n (%) | 5 (31) | 13 (16) | 0 | 0.2 |
| Post-operative stone size, mm § | 3.5 [0–9.3] | 6.0 [0.0–10.5] | 9.0 [4.5–14] | 0.5 |
| Estimated blood loss, ml | 100 [44–230] | 100 [50–200] | 150 [80–280] | 0.8 |
| Residual stones visible at the end of case, n (%) | 0 | 14 (17) | 0 | 0.1 |
| Total OR time, minutes | 98 [87–130] | 101 [79–128] | 108 [97–122] | 0.8 |
| Clavien-Dindo complication, n (%) §§: |  |  |  |  |
| Total | 4 (25) | 14 (17) | 1 (25) | 0.7 |
| II | 4 (25) | 8 (9.6) | 1 (25) | 0.2 |
| Acute Renal Failure | 0 | 1 (1.2) | 0 |  |
| Perinephric hematoma w/o emboli | 1 (6.3) | 1 (1.2) | 0 |  |
| Pneumonia | 1 (6.3) | 0 | 0 |  |
| Sepsis (No ICU) | 1 (6.3) | 4 (4.8) | 1 (25) |  |
| Blood transfusion | 1 (6.3) | 1 (1.2) | 0 |  |
| IIIa | 0 | 1 (1.2) | 0 | 0.8 |
| Hemothorax with chest tube | 0 | 1 (1.2) | 0 |  |
| IIIb | 0 | 2 (2.4) | 0 | 0.7 |
| Perinephric hematoma with emboli | 0 | 1 (1.2) | 0 |  |
| Access site fistula | 0 | 1 (1.2) | 0 |  |
| IV | 0 | 3 (3.6) | 0 | 0.5 |
| Sepsis (ICU) | 0 | 1 (1.2) | 0 |  |
| Renal artery pseudoaneurysm w embolization | 0 | 1 (1.2) | 0 |  |
| NSTEMI | 0 | 1 (1.2) | 0 |  |

Continuous variables are displayed in median [IQR].
§On operative side
§§ Within 30 days of surgery

### Intention to treat analysis

There was a concern that more complex cases were both more likely to be converted to fluoroscopy and hence at risk for lower SFR. To address this possible selection bias, an intention-to-treat analysis was performed. The SFR (Grade A, B and C) for all cases initially planned for ultrasound-guided PCNL (i.e., groups A and C) was compared to the SFR for those performed with fluoroscopy guidance (group B). No differences were seen between these two groups for either category of SFR (A, B or C) [35% vs 30%, 5% vs 1%, 15% vs 11%, in category A, B and C, respectively, p=0.6].

## Discussion

In this single-institution prospective cohort study, we demonstrate that radiation-free PCNL achieves stone-free rates (SFR) comparable to traditional fluoroscopic PCNL, supporting previous studies comparing ultrasound-only and fluoroscopy-guided PCNLs ^5,9^. More importantly, this study further adds to growing body of literature confirming the feasibility of performing a *completely* radiation-free PCNL^5,6,9–16^. However, this study does provide information distinctly different from prior reports—i.e. it also identifies cases where conversion to fluoroscopy was necessary, providing insight into technical limitations. It highlights that adoption of a radiation-free technique is not seamless. Ideally, all cases can be performed using radiation-free technique but in reality there may be barriers and understanding these obstacles may flatten the learning curve.

Of our 36 subjects that were eligible for radiation-free PCNL, 11 were converted into fluoroscopic assisted technique due to various reasons. Thematically the root cause stemmed from an overridingly similar issue (verify stent position, verify kidney anatomy, difficulty seeing dilators, difficulty seeing needle)—i.e. *poor visualization*. And this is unsurprising as ultrasonography is highly dependent on operator handling of the probe and operator interpretation of images. However, it also unmasks a gap in the field of ultrasound-based PCNL. Devices are intended for fluoroscopy—not for echogenicity. Devices created for intended acoustic energy localization require different properties yet to be well developed in endourology. Some needles are marketed as “more echogenic,” but no evidence supports their superiority over standard needles. The Boston Scientific Amplatz Super Stiff guidewire interestingly has very echogenic properties anecdotally in our series over standard guidewires.

In all, this gap in technology merits further development. Of note, we attempted to find predictors for the likelihood of conversion to fluoroscopy in initially planned radiation-free cases by performing a binominal logistic regression analysis using gender, age, ethnicity, BMI, total stone burden and prone positioning. However, none of these variables were shown to predict conversion to fluoroscopy.

Notwithstanding the inability to complete all cases with a radiation-free PCNL technique, we were able to achieve comparable results from a stone free outcome perspective. Compared to a historical series of this surgeon’s recent PCNL’s, the radiation-free PCNL achieved similar SFR’s. Admittedly SFRs in our study were lower than those reported in general populations. A 2019 meta-analysis reported SFR (Grade C) of 77.3–88% ^17^, and yet our reported SFR are consistent with the institution’s historical SFR (Grade C) of 67% ^18^.

A 2016 study found prior stone surgery and preoperative stone burden to be the main predictors of SFR in patients with staghorn calculi ^19^. While we lacked data on surgical history, we found a significant association between low preoperative stone burden and SFR on bivariate analysis (p<0.01, Table 4a). Post hoc analysis showed that lower stone burden was linked to higher SFR Grade A [p<0.05]. Male sex [p<0.05] and ASA class [p<0.05] also differed between stone-free and non–stone-free groups. Further research is needed to clarify these associations.

**Table 4a.** pre-operative differences between post-operatively stone free subjects and subjects that were not stone free post-operatively.

|  | Zero stone | Stone 0-2mm | Stone 2-4mm | Stone >4mm | p value |
| --- | --- | --- | --- | --- | --- |
| No. pts | 32 (31) | 2 (1.9) | 12 (12) | 57 (55) |  |
| Age, years | 63 [45–71] | 53 [52–53] | 56 [51–62] | 61 [51–71] | 0.4 |
| Male sex, n (%): | 15 (47) | 2 (100) | 5 (42) | 12 (21) | <0.05 |
| Ethnicity, n (%): |  |  |  |  | 0.6 |
| Caucasian | 20 (63) | 1 (50) | 7 (58) | 35 (61) |  |
| African American | 0 (0.0) | 0 (0.0) | 0 (0.0) | 2 (3.5) |  |
| Asian | 3 (9.4) | 0 (0.0) | 2 (17) | 7 (12) |  |
| Hispanic | 4 (13) | 0 (0.0) | 2 (17) | 6 (11) |  |
| Other/mixed | 4 (13) | 1 (50) | 1 (8.3) | 7 (12) |  |
| Unknown | 1 (3.1) | 0 (0.0) | 0 (0.0) | 0 (0.0) |  |
| Body mass index, kg/m<sup>2</sup> | 28 [24–31] | 27 [26–28] | 25 [24–28] | 26 [22–33] | 0.6 |
| H/o diabetes, n (%) | 10 (31) | 0 | 2 (17) | 11 (19) | 0.5 |
| H/o hypertension, n (%) | 18 (56) | 0 | 3 (25) | 32 (56) | 0.1 |
| H/o coronary artery disease, n (%) | 6 (19) | 0 | 0 | 8 (14) | 0.4 |
| H/o hyperlipidemia, n (%) | 11 (34) | 0 | 2 (17) | 18 (32) | 0.5 |
| H/o recurrent UTI's, n (%) | 6 (19) | 0 | 2 (17) | 16 (28) | 0.6 |
| Immuno-compromised, n (%) | 1 (3.1) | 0 | 0 | 0 | 0.5 |
| ASA Class 1-5, n (%): |  |  |  |  | **<0.05** |
| 1 | 2 (6.3) | 0 | 0 | 0 |  |
| 2 | 17 (53) | 2 (100) | 7 (58) | 15 (26) |  |
| 3 | 11 (34) | 0 | 5 (42) | 38 (67) |  |
| 4 | 2 (6.3) | 0 | 0 | 4 (7.0) |  |
| Laterality, n (%): |  |  |  |  | 0.2 |
| Right | 11 (34) | 2 (100) | 3 (25) | 25 (44) |  |
| Total stone burden, mm § | 22 [18–29] | 25 [24–26] | 20 [18–28] | 34 [23–48] | **<0.01** |
| Stones, n (%) |  |  |  |  | 0.9 |
| 1 | 19 (59) | 1 (50) | 8 (67) | 23 (40) |  |
| 2 | 8 (25) | 1 (50) | 2 (17) | 17 (30) |  |
| 3 | 3 (9.4) | 0 | 1 (8.3) | 8 (14) |  |
| 4 | 1 (3.1) | 0 | 1 (8.3) | 4 (7.0) |  |
| ≥5 | 1 (3.1) | 0 | 0 | 4 (7.0) |  |
| Upper pole presence, n (%) | 6 (19) | 0 | 3 (25) | 13 (23) | 0.8 |
| Mid pole stone presence, n (%) | 20 (63) | 2 (100) | 7 (58) | 44 (77) | 0.3 |
| Lower pole stone presence, n (%) | 14 (44) | 2 (100) | 5 (42) | 33 (58) | 0.3 |
| Ureter stone presence, n (%) | 4 (13) | 0 | 0 | 8 (14) | 0.5 |

§pre-operatively on operative side
Continuous variables are displayed in median [IQR]

**Table 4b.**
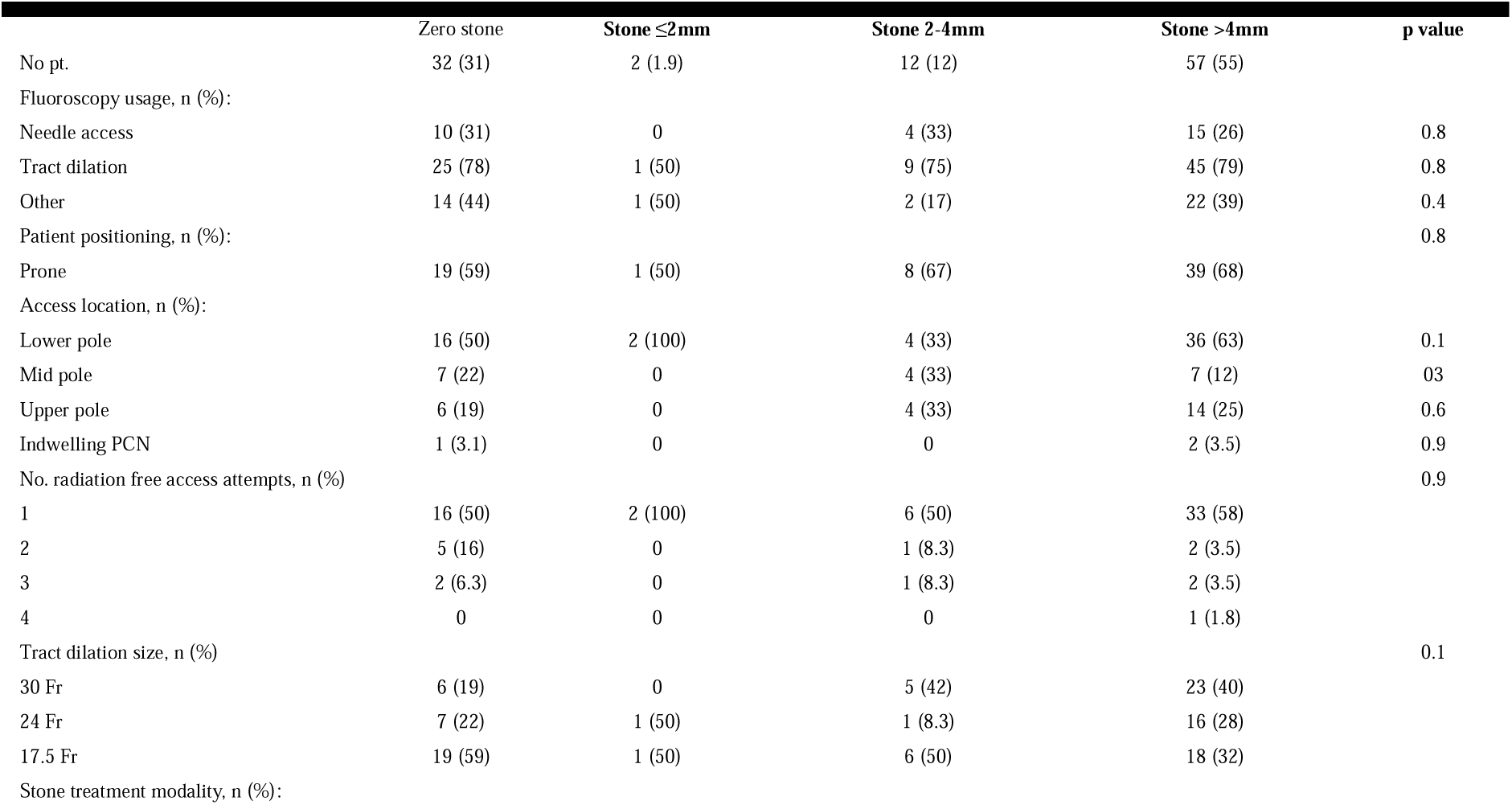
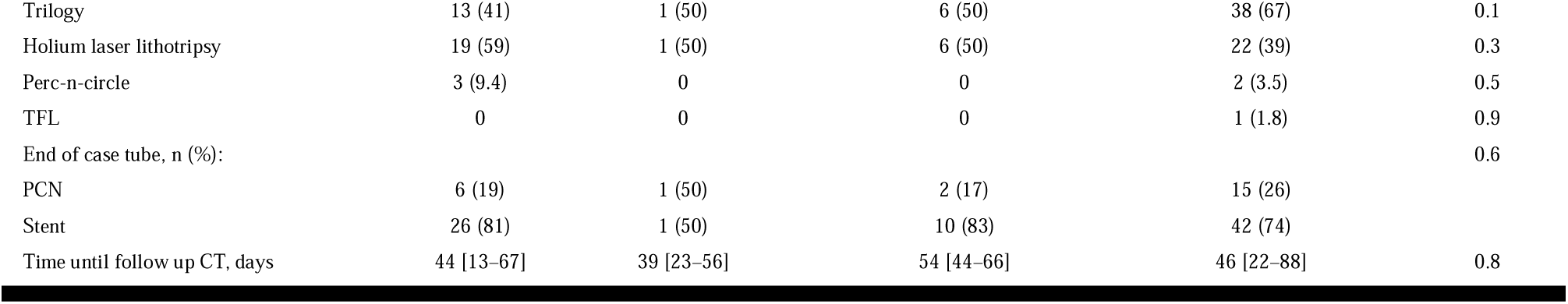
Operative differences between stone free subjects and subjects that were not stone free.

**Table 5.**
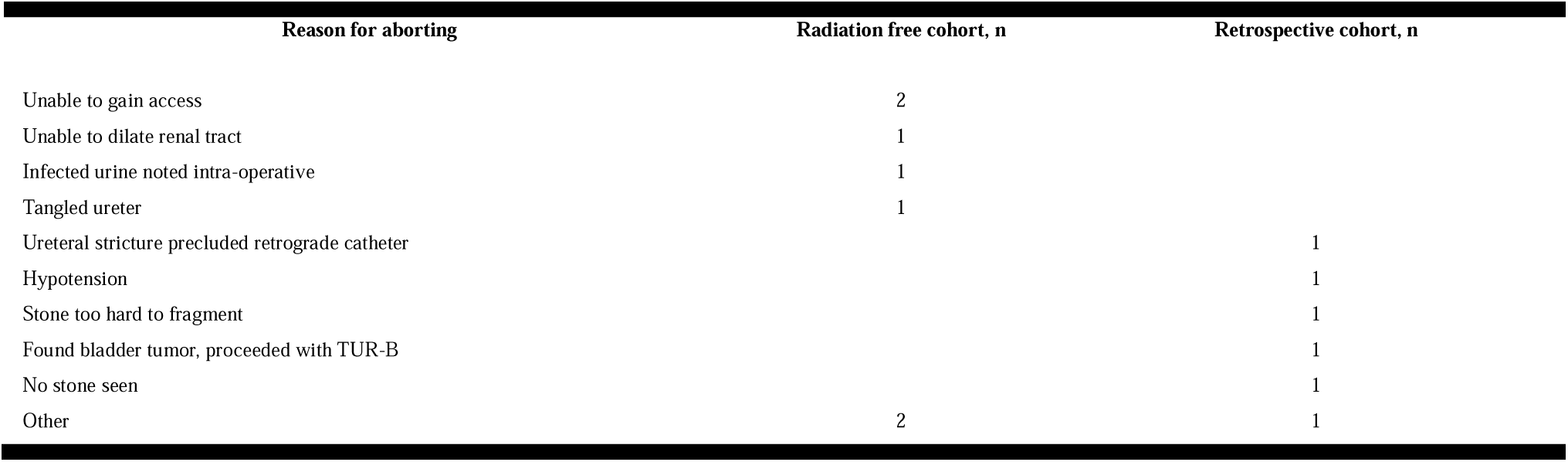
Reason for aborting PCNL.

| Reason for aborting | Radiation free cohort, n | Retrospective cohort, n |
| --- | --- | --- |
| Unable to gain access | 2 |  |
| Unable to dilate renal tract | 1 |  |
| Infected urine noted intra-operative | 1 |  |
| Tangled ureter | 1 |  |
| Ureteral stricture precluded retrograde catheter |  | 1 |
| Hypotension |  | 1 |
| Stone too hard to fragment |  | 1 |
| Found bladder tumor, proceeded with TUR-B |  | 1 |
| No stone seen |  | 1 |
| Other | 2 | 1 |

Consistent with existing literature ^5^, we observed no significant differences in secondary outcomes among Groups A, B, and C. Complication rates were 25%, 17%, and 25%, respectively, which fall within previously reported PCNL complication rates (21%–83%) ^20^. There were no significant differences in Clavien-Dindo classifications or in median estimated blood loss (EBL): 100ml in Group A, 100ml in B, and 150ml in C [p=0.8]. Comparable EBL data was not available in prior literature.

Beyond small sample size, our study has other limitations. Selection bias may be present between group A and B, as complex cases may both be associated with lower SFR and are more likely to need fluoroscopy. When comparing pre-operative stone burden between cases in a post hoc analysis, we found that subjects who were not stone free were likely to have had larger stones than subjects who were rendered stone free (Grade A). This former group also had a higher proportion of ASA 3-4 classifications, suggesting they represented more complex patients. Though we found no difference in Guy’s stone score (Table 1), which is a validated measure of complexity ^21^. In all other variables such as demographics, co-morbidities, stone location distribution, tract dilation sizes, and lithotripsy devices we did not detect differences between the cohorts.

Ultrasound’s effectiveness depends on operator experience but equally future innovation in the developing tools that are intended for ultrasound-based PCNL. In order to achieve widespread adoption of radiation-free PCNL techniques, we will likely need all PCNL products to be highly echogenic—from guidewires to dilators to sheaths to urinary drainage products.

## Conclusion

Radiation-free PCNL was safely performed with stone-free and complication rates comparable to those of fluoroscopy-guided procedures performed previously at this institution. Larger observational studies and randomized trials are needed to confirm generalizability across institutions. Future innovations focused on increasing the echogenicity of surgical products will assist the diffusion of this practice.

## Data Availability

All data produced in the present study are available upon reasonable request to the authors

## Abbreviations and acronyms

ASA: American Society of Anesthesiologists
DOAC: Direct Oral Anticoagulants
EBL: Estimated Blood Loss
IQR: Interquartile Range
OR: Operating room
PCNL: Percutaneous nephrolithotomy
SFR: Stone Free Rate
UCSD Health: University of California San Diego Health

## Acknowledgements

The authors would like to thank the operating room staff and radiology technicians at UCSD Health for their assistance during the procedures.

## Author contributions

NS: Data curation, Formal analysis, Investigation, Software, Validation, Visualization, Writing – original draft, Writing – review & editing. AG: Investigation. RH: Investigation. JG: Investigation. JR: Methodology, Investigation, Resources. JK, LG, TS: Investigation, Resources. MU: Supervision, Methodology. RLS: Conceptualization, Methodology, Supervision, Project administration, Resources.

## Supplementary information

Artificial Intelligence language tools (ChatGTP, OpenAI) were used to improve grammar and clarity of the manuscript.

